# The impact of COVID-19 pandemic on bronchiolitis (lower respiratory tract infection) due to respiratory syncytial virus: A systematic review and meta-analysis

**DOI:** 10.1101/2022.04.26.22274244

**Authors:** Sasidharanpillai Sabeena, Nagaraja Ravishankar, Sudandiradas Robin, Sabitha Sasidharan Pillai

## Abstract

**Background:** The COVID-19 pandemic has changed the epidemiology of RSV infection which accounts for most bronchiolitis cases and viral pneumonias in infants.

**Aim:** This systematic review and meta-analysis aimed to quantitatively assess the effect of COVID-19 pandemic on respiratory syncytial virus (RSV) associated bronchiolitis among hospitalised infants globally.

**Methods:** The study protocol was registered in the PROSPERO database (CRD42022314000) and was designed based on PRISMA guidelines updated in May 2020. An electronic search of PubMed/MEDLINE, Scopus and Google Scholar was carried out for articles regarding the impact of the COVID-19 pandemic on bronchiolitis or lower respiratory tract infection due to the respiratory syncytial virus in English published between January 2019 and March 2022. The meta-analysis component was modified appropriately to synthesise the pooled proportion of infants having RSV-associated bronchiolitis before the COVID-19 pandemic in 2019 and during the pandemic with 95% confidence interval (CI).

**Results:** We screened 189 articles and systematically reviewed fifty studies reporting RSV-associated bronchiolitis cases in infants before the pandemic in 2019 and during the pandemic in 2020/2021. Eight qualified studies from Europe and China, which reported RSV-bronchiolitis both in 2019 and in 2020/21 were pooled by random-effects meta-analysis. These studies comprised 109,186 symptomatic cases of bronchiolitis before the pandemic in 2019 and 61,982 cases in 2020-2021. The quantitative analysis included laboratory-confirmed RSV infection in 7691 infants with bronchiolitis reported before the pandemic in 2019. Meanwhile, during the pandemic, 4964 bronchiolitis cases were associated with RSV infection. The pooled proportion of RSV-associated bronchiolitis cases before the pandemic in 2019 was 16.74% (95% CI 11.73, 22.43%, 95% prediction interval 0.032, 34.16). The pooled proportion of confirmed RSV cases during the pandemic in 2020/2021 was 19.20 % (95% CI 12.01, 27.59%, 95% prediction interval 0.046, 42.35).

**Conclusion:** There was an increase in RSV activity after the relaxation of stringent public health measures during the COVID-19 pandemic.

**Key Messages (Provide appropriate messages of about 35-50 words to be printed in centre box):**

- This systematic review and meta-analysis reports the pooled proportion of RSV associated bronchiolitis cases in 2019 (before the COVID-19 pandemic) and during the pandemic.
- Eight observational studies from China and Europe were qualified for the meta-analysis.
- A decline in reported cases of bronchiolitis was observed during the COVID-19 pandemic which might be attributed to non-pharmaceutical measures and a fall in the hospitalisation rates of respiratory non-SARS-CoV-2 infections.
- The pooled proportion of RSV positivity rate among bronchiolitis cases was more during the COVID-19 pandemic.

## Introduction

Severe acute respiratory syndrome coronavirus 2 (SARS-CoV-2) is a highly infectious novel coronavirus that emerged in late 2019, leading to the coronavirus disease 2019 (COVID-19) pandemic. The non-pharmaceutical interventions such as masking, use of hand sanitisers and various public health measures of lockdowns, travel restrictions, social distancing, isolation, and quarantines helped to control the transmission in the initial months. Compared to the pre-pandemic seasons, the epidemiology of other common viral respiratory infections was significantly affected during the pandemic ^1^. There was a decrease in respiratory viral infections, including respiratory syncytial virus (RSV), due to various non-pharmaceutical interventions implemented to curb the pandemic. These public health measures reduced the transmission of enveloped viruses such as influenza and RSV ^2^. Lower respiratory tract infection or bronchiolitis in infants leads to increased hospitalisations and mortality during the winter season. The primary aetiology of paediatric bronchiolitis is RSV infection, which results in recurrent wheeze and asthma in childhood ^3^. Most of the bronchiolitis cases occur in low-income countries with limited RSV surveillance, which demands molecular diagnostic facilities. Over the past year, not a single RSV case was reported in many countries during the winter season, which extended between the last months of 2020 and early 2021. Many developed countries in the northern and southern hemispheres reported an unexpected surge in the RSV cases during the summer of 2021^4^.

COVID-19 pandemic has led to alterations in the epidemiology of RSV infection which accounts for most bronchiolitis and viral pneumonias in infants. During the spring or summer seasons, a resurgence in areas reporting a meagre number of RSV cases was noted globally during the pandemic. Monitoring the effect of COVID-19 on the circulation of RSV infection in infants is essential as there are no specific vaccines or antivirals for this virus. This systematic review and meta-analysis aimed to analyse the effect of the COVID-19 pandemic on RSV-associated bronchiolitis among hospitalised infants.

## Methods

We started the systematic review after excluding registered or ongoing systematic reviews regarding the impact of the COVID-19 pandemic on bronchiolitis (lower respiratory tract infection) due to respiratory syncytial virus in the PROSPERO database. The study protocol was registered in the PROSPERO database (CRD42022314000) and can be accessed at https://www.crd.york.ac.uk/prospero/display_record.php?ID=CRD42022314000.

The systematic review and meta-analysis were performed based on the Preferred Reporting Items for Systematic Reviews and Meta-analyses (PRISMA) guidelines updated in May 2020^5^. The meta-analysis component was modified appropriately to synthesise the pooled proportion of paediatric bronchiolitis cases tested positive for RSV before the COVID-19 pandemic in 2019 and during the pandemic.

## Description of the condition

### Bronchiolitis

Bronchiolitis is the most common lower respiratory tract viral infection caused by RSV among children below two years ^6^, followed by rhinovirus and human Boca virus. The main clinical features of bronchiolitis are cough, chest recession or tachypnoea, crackles on chest auscultation or the first episode of acute wheeze ^7^.

### Extended severe acute respiratory infection (SARI) due to RSV

Symptoms of cough/shortness of breath within the last ten days necessitating hospitalisation due to RSV is defined as extended severe respiratory infection due to RSV ^8^.

### RSV infection

RSV infection was confirmed in symptomatic cases by real-time reverse transcription polymerase chain reaction (RT-PCR), immunofluorescence assay or virus isolation.

#### 2 Study Protocol

An electronic search of PubMed/MEDLINE, Scopus and Google Scholar was carried out for all the articles in English published between January 2019 and March 2022 regarding the impact of the COVID-19 pandemic on bronchiolitis or lower respiratory tract infection due to the respiratory syncytial virus using search terms such as “impact” AND “COVID-19” OR “SARS-CoV-2” AND “RSV” NOT “vaccines”. Manual library search for articles published in peer-reviewed journals was carried out. The references of retrieved articles were examined to increase the search sensitivity.

#### 3 Inclusion process and criteria

Observational studies in English reporting the number of immunocompetent paediatric cases aged two years or below having bronchiolitis or lower respiratory tract infection due to RSV infection were included. Conference abstracts and commentaries were excluded. The studies reporting the data among older children and adults were eliminated. The articles reporting the RSV activity among immunocompromised cases were also not considered.

#### 4 Data extraction

A validated proforma detailing the name of the first author, year of publication, study area, number of cases with bronchiolitis/lower respiratory tract infection (LRTI) attended the hospitals in 2019 and during the pandemic, number of laboratory-confirmed RSV positive cases in 2019 and during the pandemic 2020/2021 was prepared. A three-stage selection of published articles was carried out for the final inclusion. One reviewer evaluated the titles of the records for the relevance for inclusion in the study (n=5479). Studies applicable for the review were moved to the second stage after eliminating irrelevant topics and duplicates (n=5290). In the second stage, the abstracts of the studies were obtained and were independently examined by two reviewers (n=97). After reviewing the abstracts, full texts of studies were procured, which were inspected by two reviewers independently (n=50). The corresponding authors were communicated electronically if further clarification was needed. Manual library search for articles in peer-reviewed journals was carried out, and references of retrieved articles were reviewed to increase the search sensitivity. The PRISMA 2020 flow diagram ^5^ (**Figure 1**) depicts the study selection process. The last date of the search was on March 22, 2022.

**Figure 1.**
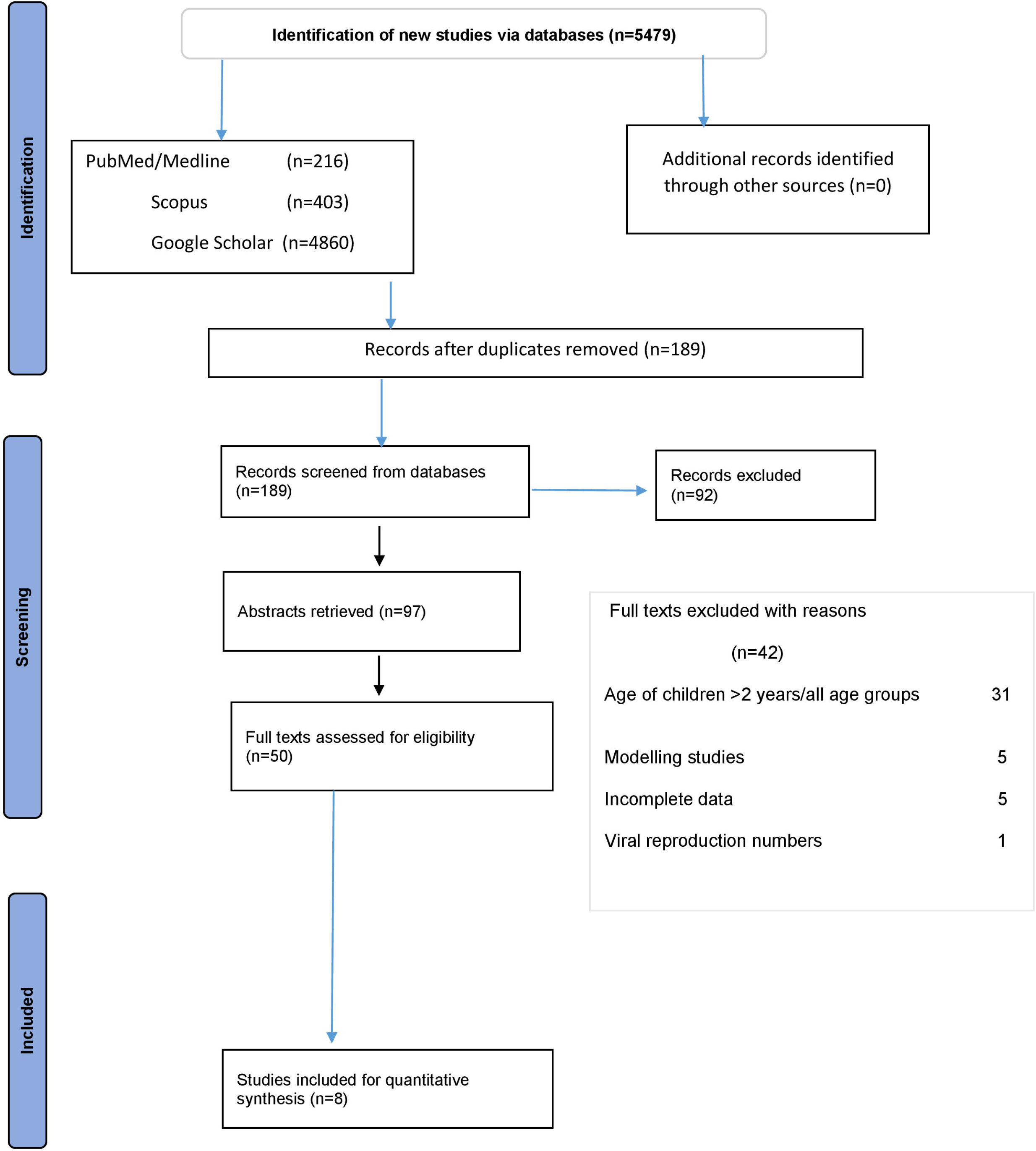
The PRISMA 2020 flow diagram for updated systematic reviews which included searches of databases, registers and other sources^5^. The flow diagram illustrates the number of studies identified, screened, abstracts/full-text articles included/excluded for the systematic review and meta-analysis.

#### 5 Risk of bias (quality) assessment in individual studies

To assess the risk of bias in individual studies (quality assessment), chosen after the abstract and content review, the National Institutes of Health checklist for observational, cohort and cross-sectional studies was used^9^. The studies with a minimum score of eight or above, seven, or five or less than five “Yes responses” were considered good, fair, and poor quality, respectively. For cross-sectional studies, question numbers 1, 2, 3, 4, 5 and 11 were applicable. The responses to the remaining eight questions (6-10, 12, 13, 14) were not applicable (NA). The studies with six “Yes” responses were considered good, and those with four /five were taken as fair. The studies with less than four “Yes responses” were considered poor quality. The quality of the studies was evaluated by two reviewers independently.

#### 6 Statistical analysis

The meta-analysis was accomplished in STATA version 13.0 (College Station, Texas 77, 845 USA). The forest plots were constructed using metaprop package in STATA. The pooled proportion of symptomatic cases of RSV-associated bronchiolitis or lower respiratory tract infections before the pandemic in 2019 and during the pandemic was reported with 95% confidence interval (CI). Chi-square statistic (Q statistic) and I^2^ index to quantify the heterogeneity was noted as considerable heterogeneity across the studies was expected. The I^2^ value ranging between 0% to 24% specifies consistency. I^2^ values of 25%-49% imply low heterogeneity and 50-74% points toward moderate heterogeneity. The I^2^ value varying between 75%-100% is indicative of high heterogeneity^10^.

#### 7 Prediction interval

The concept of prediction interval was introduced in 2016 to address the uncertainty in the summary effect if a new study was incorporated in the meta-analysis ^11^. The precision of the mean effect size is measured by the CI. Meanwhile, the prediction interval indicates the dispersion of the true effect size of the newly added studies ^12^. In the forest plot, centre of the diamond depicts the summary effect size and the horizontal line to either side illustrates the prediction interval. Prediction interval facilitates the clinical interpretation of the heterogeneity.

#### 8 Assessment of Publication bias

Egger’s test was used to report the publication bias. Weighted linear regression with standardised effect estimate and precision was considered the dependent and independent variables, respectively. In the present study, the log_e_ proportion of RSV positive bronchiolitis cases and precision were considered the effect estimate and 1/standard error of log_e_ proportion rate, respectively. Weights were allotted using the inverse variance approach (1/variance of the effect estimate). The statistically significant bias coefficient implies publication bias (*p*-value<0.05).

## Results

### Included studies

We screened 189 articles using the search terms and ninety-seven abstracts were retrieved. Fifty articles reporting RSV-associated bronchiolitis cases in infants before the pandemic in 2019 and during the pandemic in 2020/2021 were systematically reviewed. Eight studies qualified for the meta-analysis were from Spain, Italy, France and China, including 109,186 cases of bronchiolitis before the pandemic in 2019 and 61,982 cases during the pandemic^13–20^. All these studies were qualified as good as shown in **Table 1**. These studies reported 7691 laboratory-confirmed RSV associated lower respiratory tract infections in 2019 and 4964 during the pandemic in 2020/21. Two studies from Spain and France were multicentric ^18,20^. The highest number of symptomatic cases were admitted in the multicentric study from Spain before and during the pandemic ^18^. All the studies chosen for the meta-analysis reported a decrease in bronchiolitis cases with increased RSV positivity rate during the pandemic. The highest rate of RSV positivity of 70.2% in bronchiolitis was noted from Spain just before the pandemic in 2019. Meanwhile, during the pandemic RSV was the causative agent in up to 77.8% of bronchiolitis cases from the same study area. The number of infants admitted were the lowest in the above mentioned study from Barcelona, Spain. Another large multicentric study from Spain reported RSV positivity of 7.2% before the pandemic and 9.1% during the pandemic^13^. Both the studies from China and multicentric study from Spain included cases seen during the entire year 2019 and whole 2020. Meanwhile, single centre studies from Barcelona, Italy and France enrolled infants with bronchiolitis admitted during the timeframe spanning over eight to ten months. The lowest RSV positivity in bronchiolitis cases was noted in a multicentric study from Lyon, France before and during the pandemic as in the Table 1. Before the pandemic in 2019, the study from Lyon reported 124 (1.35%) laboratory confirmed RSV cases out of 9,127 infants during the RSV season extending from September to May. During the corresponding months in 2020, seventy-seven RSV positive bronchiolitis cases were reported out of 8728 symptomatic infants hospitalised with a positivity rate of 0.88% ^20^. Both the studies from China enrolled infants below the age of 12 months and RSV infection was confirmed by multiplex direct IFA. The study from Shanghai reported RSV positivity rate of 6.6% among 4600 hospitalised infants in 2019, which declined to 4.5% in 2020 when 2507 bronchiolitis cases were admitted ^16^. Another study from Hangzhou region observed a rise in the RSV positivity in 2020 compared to 2019, even though the number of admitted infants was less ^19^.

**Table 1:** The table depicts the characteristics of the study included for the meta-analysis.

| S<br>N<br>o | Reference<br>(year) | Region | Study<br>design | Age<br>(months) | *LRTI/<br>bronchiolitis<br>before the<br>pandemic | Duration | †RSV<br>Positivity<br>(N) | Percentage<br>(%) | *LRTI/<br>bronchiolitis<br>during the<br>pandemic | Duration | †RSV<br>Positivity<br>(N) | Percentage<br>(%) | Diagnosis | Quality |
| --- | --- | --- | --- | --- | --- | --- | --- | --- | --- | --- | --- | --- | --- | --- |
| 1 | Guitart et al<br>2022<sup>13</sup> | Barcelona<br>Spain | Prospective<br>observational | <12 | 124 | 2019 Sept-June 2020 | 87<br>(70.2) | 70.2 | 63 | 2020<br>Sept-June<br>2021 | 49 | 77.8 | RT-PCR‡ | Good |
| 2 | Stera et al<sup>14</sup><br>2021 | Bologna<br>Italy | retrospective<br>observational | ≤12 | 114 | Oct 2018-April 2019 | 75<br>(65.8) | 65.8 | 98 | 2019/20<br>20<br>Oct-April | 72 | 73.5 | RT-PCR | Good |
| 3 | Curatola et<br>al 2021<sup>15</sup> | Rome<br>Italy | retrospective<br>observational | ≤24 | 238 | 2019 Feb-2020 Feb | 40<br>(16.8) | 16.8 | 33 | 2020<br>Feb-2021 Feb | 6 | 18.2 | RT-PCR | Good |
| 4 | Liu et al<br>Aug 2021<sup>16</sup> | Shanghai<br>China | retrospective<br>observational | ≤12 | 4600 | 2019<br>Jan-Dec | 304<br>(6.6) | 6.6 | 2507 | 2020<br>Jan-Dec | 114 | 4.5 | Multiplex<br>Direct<br>IFA § | Good |
| 5 | Fourgeaud<br>et al 2021<sup>17</sup> | France | retrospective<br>observational | ≤11 | 5396 | Aug 2019-July 2020 | 172<br>(3.2) | 3.2 | 4804 | 2020/21-Aug-April | 169 | 3.5 | RT-PCR | Good |
| 6 | Torres-Fernandes<br>et al 2021<sup>18</sup> | Spain | retrospective<br>observational | ≤24 | 82,577 | 2019<br>Jan-Dec | 5936<br>(7.2) | 7.2 | 42,924 | 2020<br>Jan-Dec | 3912 | 9.1 | RT-PCR | Good |
| 7 | Ye et al<sup>19</sup> | Hangzhou<br>China | retrospective<br>observational | ≤12 | 7010 | 2019<br>Jan-Dec | 953<br>(13.6) | 13.6 | 2825 | 2020<br>Jan-Dec | 565 | 20.0 | Multiplex<br>Direct<br>IFA | Good |
| 8 | Casalegno et<br>al 2021<sup>20</sup> | Lyon<br>France | retrospective<br>observational | ≤12 | 9,127 | 2019/20<br>Sept-May | 124<br>(1.35) | 1.35 | 8,728 | 2020/21<br>Sept-May | 77 | 0.88 | RT-PCR | Good |

\* LRTI lower respiratory tract infection
†RSV respiratory syncytial virus
‡RT-PCR reverse transcription polymerase chain reaction
§IFA immunofluorescence assay

### Meta-analysis

The pooled proportion of RSV-associated bronchiolitis cases before the pandemic in 2019 was 16.74% (95% CI 11.73%,22.43%) as in **Figure 2**. The **Figure 3** represents the pooled proportion of RSV cases among infants during the pandemic as 19.20% (95% CI 12.01%,27.59%). As the I^2-^value was>90%, a high heterogeneity was observed between the studies.

**Figure 2.**
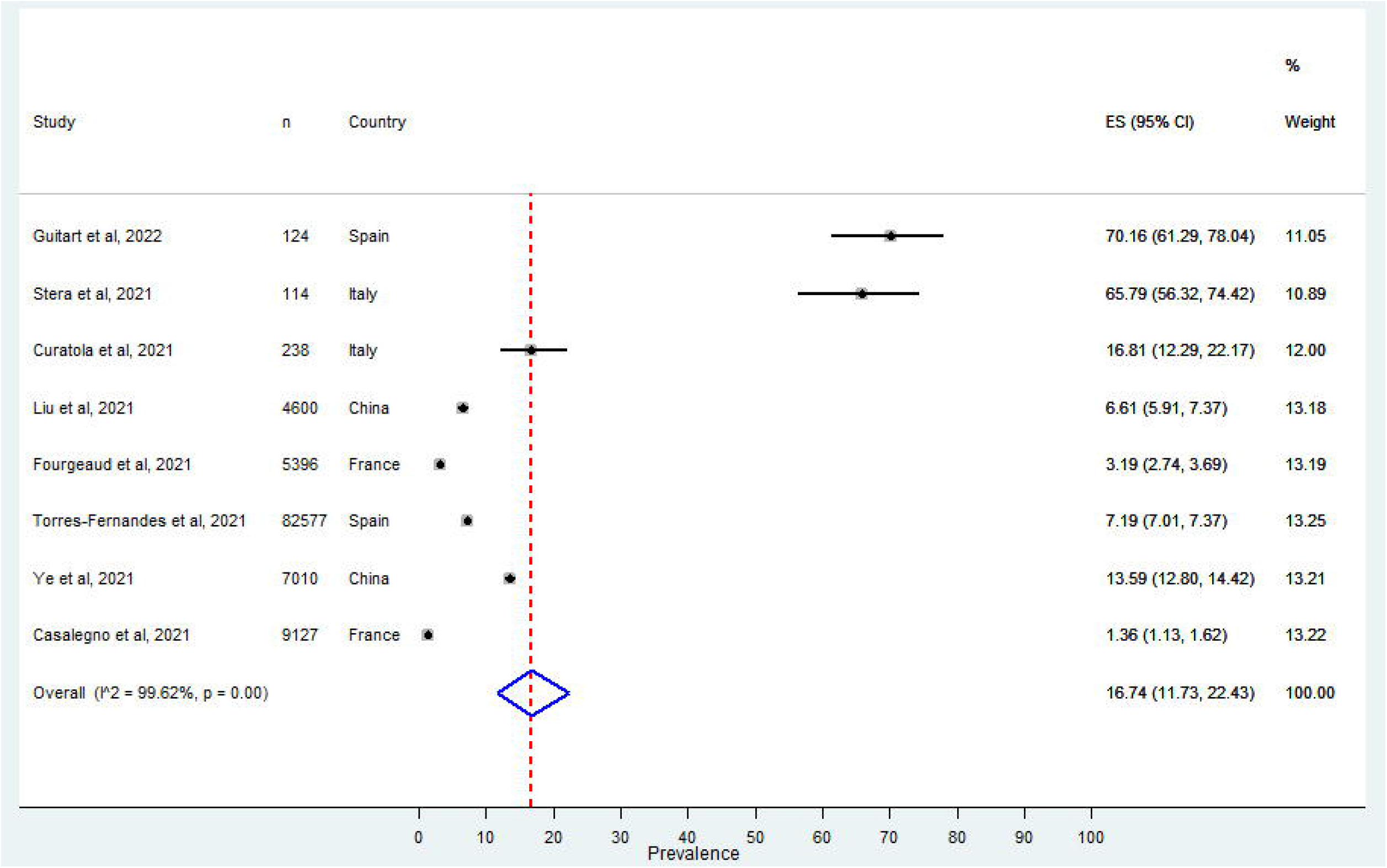
The Forest Plot of the summary effect size (Proportion of bronchiolitis cases tested positive for RSV before the COVID-19 pandemic) using random-effects model. Squares indicate the effect size of individual studies and the extended lines denote 95% confidence intervals (CI). Sizes of squares imply the weight of studies based on sample size using a random effects analysis. The diamond data indicates pooled proportion. Test of heterogeneity: I^2=^ 99.62%, *p*-value=0.00.

**Figure 3.**
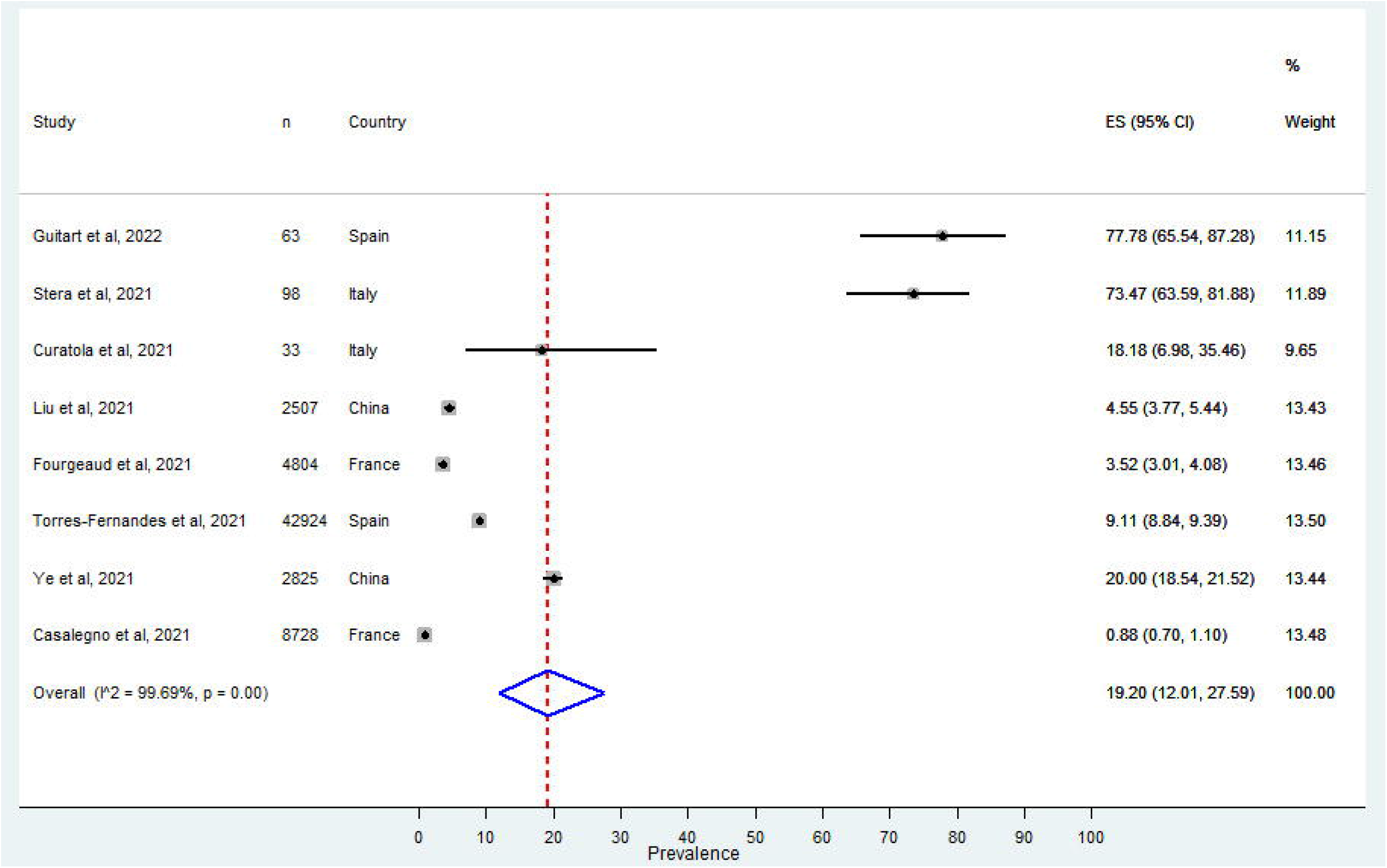
The Forest Plot of the summary effect size (Proportion of RSV positive bronchiolitis cases during the COVID-19 pandemic) using random-effects model. Squares indicate the effect size of individual studies and the extended lines denote 95% confidence intervals (CI). Sizes of squares imply the weight of studies based on sample size using a random effects analysis. The diamond data indicates pooled proportion. Test of heterogeneity: I^2=^ 99.69 %, *p*=0.00.

### Prediction interval

The 95% prediction interval was 0.032, 34.16 before the pandemic in 2019 and 0.046, 42.35 during the pandemic.

### Publication bias

As shown in **Table 2**, the *p*-value for the bias coefficient was not statistically significant. Hence there was no publication bias.

**Table 2:** The table depicts the results of Egger’s test for publication bias.

| **Results of Egger's test (Before the Pandemic)** |  |  |
| --- | --- | --- |
| Coefficient | Estimate (95%CI) | P-value |
| Slope | 0.0532 (0.0067, 0.0997) | 0.051 |
| Bias | 2.9071 (-2.5261, 8.3403) | 0.238 |
| **Results of Egger's test (after the Pandemic)** |  |  |
| Slope | 0.0605 (-0.0093, 0.1304) | 0.449 |
| Bias | 2.4388 (-3.7147, 8.5924) | 0.370 |

## Discussion

The studies qualified for this meta-analysis were from China and Europe. The pooled proportion of RSV positivity in bronchiolitis cases was more during the COVID-19 pandemic. The decline in hospitalised bronchiolitis cases or lower respiratory tract infections during the COVID-19 pandemic might be attributed to reduced admission rates for non-SARS-CoV-2 respiratory infections during the pandemic. Many countries such as Italy, Germany, Belgium, Vienna and Brazil reported no hospitalised bronchiolitis cases during the winter season ^14,21–23^. Similarly, the study from Israel reported a temporal shift of RSV epidemic to spring season from winter ^24^. During the COVID-19 pandemic, hospitals in Spain reported a steep decline in bronchiolitis cases and RSV positivity with seven months delayed RSV peak in July 2021^18^. Meanwhile, France reported a rise in RSV cases during the late winter months of 2021 after the second national lockdown in December 2020.^17^ The tropical country of Thailand, located in South East Asia, witnessed a delayed RSV season in 2020 among children with influenza-like illness^25^. There was an increase in the number of confirmed RSV cases across all age groups during the spring season of the year 2020 in Australia and South Africa ^26,27^. The inter-seasonal rise in RSV infections during the summer was observed in the northern and southern hemispheres ^28,29^. Italy reported a significant decline in hospitalised cases of bronchiolitis in 2020, with no bronchiolitis cases in 2021. However, a late RSV peak was not observed in Italy, unlike many countries ^14^. In 2020, China reported a decrease in lower respiratory tract infection among children, mainly due to the strict public health measures to contain the SARS-CoV-2 infection. The studies from Shanghai and Xiamen reported a decline in RSV activity among the hospitalised children. Both the studies observed a mild increase in RSV and influenza activities during the winter months without a temporal shift in seasonality^16,30^. However, another single centre study from Hangzhou reported a marked rise in RSV cases during the winter season of 2020, comparable to the pre-pandemic period ^19^.

Respiratory viral infections at young age facilitate the building up of trained immunity. Apart from a relatively young population of Africa compared to other parts of the world, prior exposure to other cross-reactive viruses is another causative factor for low SARS-CoV-2 related morbidity and mortality in Sub-Saharan Africa^31^. However, countries perceiving a very low number of RSV cases are at an increased risk of resurgence of RSV infection, mainly among older RSV-naïve children^32^. Sentinel surveillance of RSV is better in many European countries than in other parts of the world ^33,34^. There is no data regarding the RSV activity from the majority of the Asian and African countries having higher RSV associated disease burden ^35^.

### Causes of heterogeneity

The main heterogeneity was in the number of bronchiolitis cases enrolled before and during the pandemic. Two multicentric studies enrolled a higher number of hospitalised cases compared to single centre studies. Even though six studies confirmed RSV infection by real time reverse transcription polymerase chain reaction (RT-PCR), two studies from China detected the RSV infection by direct immunofluorescence assay^16,19^. Immunofluorescence assay is a cost-effective, reliable point-of-care assay for the diagnosis of RSV infection with less sensitivity compared to PCR ^36^. Another disparity was in the time frame during which the samples were procured from paediatric bronchiolitis cases. Although six studies enrolled infants below one year, two studies enrolled cases below two years.

### Strengths

The catchment population (denominator) was well defined. Corresponding authors were contacted electronically for retrieval of complete data. The present meta-analysis reports 95% prediction interval along with confidence interval for more explanatory clarification.

### Limitation

Major limitation was the heterogeneity of studies pooled for quantitative synthesis. Before the COVID-19 pandemic, there was no routine testing of bronchiolitis samples for RSV from developing countries in South East Asia, Africa and Latin America. The studies incorporated in the meta-analysis were from Europe and China. Even though there were studies from other parts of the world reporting low RSV activity among children during the COVID-19 pandemic, very few studies reported the RSV positivity rate among symptomatic cases. The studies pertaining to RSV surveillance from the US, Germany and South Africa could not be included for the meta-analysis due to lack of data among infants ^27,37–40^. The change in priorities and economic constraints might have led to reduced testing for non-SARS-CoV-2-respiratory viruses in developing countries ^41^. Even though the search was limited to the studies published in English, no relevant studies were omitted. All the studies reporting RSV-associated bronchiolitis among infants were hospital based.

Except France and Iceland reporting delayed outbreaks, European countries reported no RSV outbreaks since the implementation of non-pharmacological measures to control the COVID-19 pandemic ^42^. Uninterrupted RSV surveillance in coming years is vital in Europe as larger outbreaks beyond the previously reported RSV seasonality is anticipated. During the coming RSV seasons, health care professionals and hospitals must be organised for the timely detection and management of the increasing number of admissions. This systematic review will add value to the European Union funded PROMISE project initiated in November 2021 which aims RSV surveillance and immunisation in Europe ^43^.

## Conclusion

There was an increase in RSV activity after relaxing stringent public health measures during the COVID-19 pandemic in the northern and southern hemispheres. In spite of observing low number of bronchiolitis cases, the number of RSV associated paediatric bronchiolitis cases was more during the COVID-19 pandemic. Efforts for the diagnosis and surveillance for RSV must be reinforced along with influenza and SARS-CoV-2 viruses to lessen the burden of morbidity and mortality due to RSV-associated bronchiolitis among infants.

## Data Availability

All data produced in the present study are available upon reasonable request to the corresponding author.

https://www.crd.york.ac.uk/prospero/display_record.php?ID=CRD42022314000

## Notes

### Competing Interest Statement

The authors have declared no competing interest.

### Funding Statement

This study did not receive any funding.

### Summary of Updates

The abstract of the article was modified.

